# Adjunctive Role of SGLT2 Inhibitors in Transthyretin Cardiac Amyloidosis: A Systematic Review and Meta-Analysis

**DOI:** 10.64898/2026.01.14.25343185

**Authors:** Victor Lopez, Melissa Chacon, Mahmoud Abdalla, Augustine Agocha

## Abstract

**Background:** Transthyretin amyloid cardiomyopathy (ATTR-CM) has historically been underdiagnosed but has recently become increasingly recognized due to advances in diagnostic techniques and heightened clinical awareness. Despite this progress, treatment options remain limited, as current approved therapies are costly and not widely accessible. Given the benefits of sodium-glucose cotransporter 2 (SGLT2) inhibitors in broader heart failure (HF) populations, we aimed to evaluate their efficacy in reducing mortality and hospitalizations in ATTR-CM.

**Objectives:** To determine whether SGLT2 inhibitors reduce all-cause mortality, CV mortality, and HF hospitalizations in ATTR-CM, offering a potential adjunctive therapy for this undertreated population.

**Methods:** We performed a systematic review and meta-analysis of SGLT2 inhibitors against SGLT2 inhibitors-naïve patients with ATTR-CM. PubMed, Embase, Scopus and Cochrane databases were searched for trials published up to January 31, 2025. Data were extracted from published reports, and quality assessment was performed per Cochrane recommendations. Risk ratios (RRs) with 95% confidence interval (CI) were pooled across trials. Outcomes included all-cause mortality, CV mortality and HF hospitalization.

**Results:** Out of 177 database results, four observational studies and 5039 patients were included; 2489 (49.39%) received a SGLT2 inhibitor. All-cause mortality (RR 0.44; 95% CI 0.33-0.59; p<0.00001; I²=54%) and CV mortality (RR 0.30; 95% CI 0.16-0.55; p=0.0001; I²=25%) were significantly lower in patients treated with SGLT2 inhibitors compared with control. HF hospitalization (RR 0.68; 95% CI 0.33-1.41; p=0.30; I²=89%) showed a downward trend, yet this was not statistically significant.

**Conclusions:** In patients with ATTR-CM, SGLT2 inhibitors significantly reduce both all-cause and cardiovascular mortality compared to standard care, suggesting they may serve as a valuable adjunctive therapy for this undertreated population. Although HF hospitalization showed a nonsignificant downward trend, these findings underscore the need for large randomized trials to confirm and expand on these promising results.

## 1. Introduction

Transthyretin amyloid cardiomyopathy (ATTR-CM), whether acquired or hereditary, is characterized by the deposition of misfolded transthyretin proteins in the myocardium, leading to the formation of amyloid fibrils and progressive symptomatic heart failure (HF).^1–2^ Although historically considered rare, the past decade has seen a marked increase in ATTR-CM diagnoses, driven by advances in non-invasive diagnostic techniques and greater clinical awareness.^1–2^ These diagnostic improvements have also paved the way for the development of promising disease-modifying therapies.^2–3^ At present, the US Food and Drug Administration (FDA) has approved two medications for treating ATTR-CM—,tafamidis and acoramidis,—which have been shown to reduce cardiovascular (CV) related hospitalizations and mortality.^3–4^ However, the high cost of these treatments has limited their widespread use, resulting in supportive care remaining the mainstay of management in many regions.^5^ Moreover, there is a paucity of data regarding the effectiveness of conventional heart failure medications in ATTR-CM patients, although sodium-glucose cotransporter type two (SGLT2) inhibitors appear to be a particularly promising drug class for further investigation.^6^

SGLT2 inhibitors have demonstrated efficacy across the full range of left ventricle ejection fractions (LVEF), functioning as multifaceted agents that improve cardiac performance, reduce HF hospitalizations, and slow the progression of chronic kidney disease (CKD).^7^ ATTR-CM, an infiltrative disease that typically presents as HF with preserved EF (HFpEF), has been notably absent from SGLT2 inhibitor clinical trials, leaving their effectiveness in this patient population undetermined.^6–7^ While conventional heart failure treatments, such as beta-blockers and angiotensin inhibitors, have been associated with lower all-cause mortality in retrospective studies, their use is often limited by hemodynamic side effects as cautioned by the 2023 ATTR-CM guidelines.^8^ In contrast, SGLT2 inhibitors do not significantly impact hemodynamics offering a promising alternative for patients with ATTR-CM who have limited therapeutic options.^6–8^ Preliminary cohort studies suggest that SGLT2 inhibitors are safe and well tolerated in this setting, although further robust evidence is required to confirm their benefits in this specific population.

No randomized controlled trials (RCTs) have been conducted to assess SGLT2 inhibitors therapy in ATTR-CM patients. A recent meta-analysis by Karakasis and colleagues, which pooled data from five studies totaling 9,766 patients, represents a significant effort to consolidate the available evidence.^9^ However, we noted that three of these studies likely involved overlapping patient populations—each drawing data from the TriNetX database during similar periods—which may have led to an overestimation of the overall statistical power. To address this limitation, our analysis carefully accounted for potential population overlaps and additionally included a study of 176 patients that was omitted in the previous meta-analysis. Our aim was to perform a systematic review and meta-analysis examining the efficacy of SGLT2 inhibitors in ATTR-CM patients, with a particular focus on mortality and hospitalization endpoints.

## 2. Methodology

This systematic review and meta-analysis was conducted using the Cochrane Collaboration Handbook for Systematic Reviews of Interventions^10^ and the Preferred Reporting Items for Systematic Reviews and Meta-Analyses (PRISMA) guidelines.^11^ The study protocol was prospectively registered in the International Prospective Register of Systematic Reviews (PROSPERO), under the registration number CRD420251015355.

### 2.1 Search Strategy

We performed a comprehensive search of PubMed, Embase, Scopus, and the Cochrane Central Register of Controlled Trials from their inception through January 31, 2025, using key terms such as “*transthyretin cardiac amyloidosis*”, “*ATTR-CA*”, “*sodium-glucose cotransporter-2 inhibitors*”, “*SGLT2i*”, “*dapagliflozin*”, “*canagliflozin*”, “*empagliflozin*”, “*ertugliflozin*”, and “*bexagliflozin*”. Additionally, we screened the references of the included studies and relevant systematic reviews to identify further pertinent research.

### 2.2 Eligibility Criteria and Outcomes of Interest

Studies were eligible for inclusion if they met all of the following criteria: (1) they were either randomized trials or nonrandomized cohort studies; (2) they compared outcomes between patients treated with SGLT2 inhibitors and those who were SGLT2 inhibitors-naïve; (3) they involved patients diagnosed with ATTR-CM; and (4) they reported on the clinical outcomes of interest. Studies were excluded if they lacked a control group or if there was overlap in patient populations. In such cases, only the study with the largest patient cohort was selected. The primary endpoint for our analysis was all-cause mortality, while secondary outcomes included CV mortality and HF hospitalization.

### 2.3 Data Extraction

Two authors (V.L. and M.C.) independently extracted the baseline characteristics (shown in Table 1) and outcome data according to predetermined criteria for the literature search, data extraction, and quality assessment. Any discrepancies between them were resolved by mutual consensus.

**Table 1.**
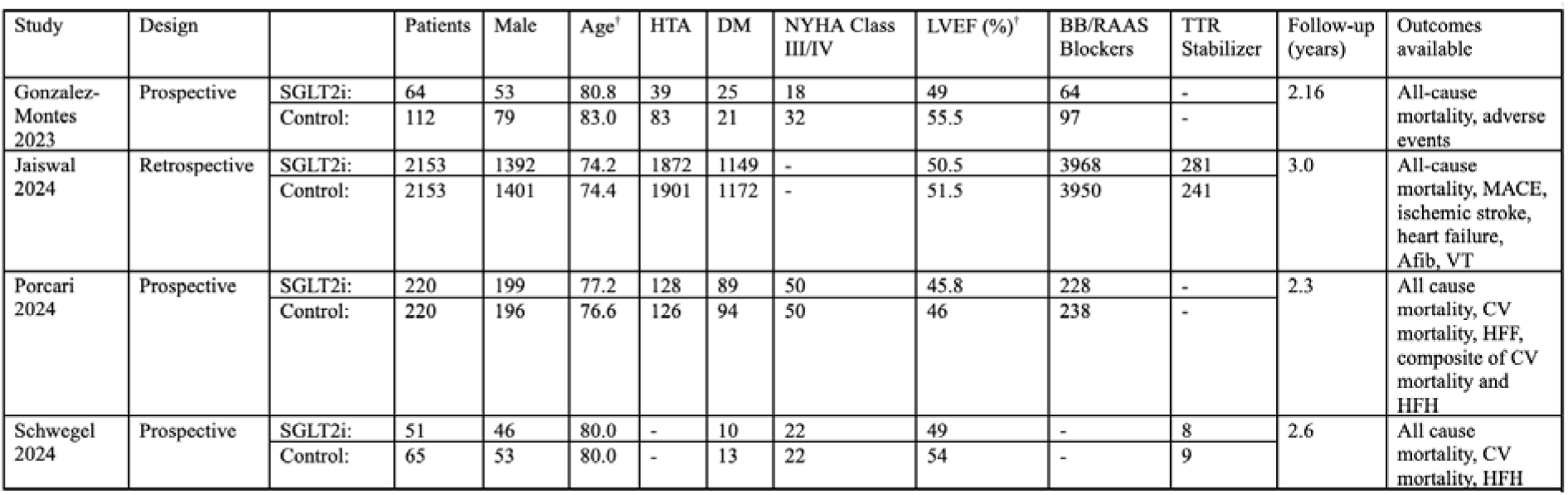
Baseline Characteristics of included studies. ^†^mean or median; SGLT2i: SGLT2 Inhibitors; HTA: Hypertension; DM: Diabetes Mellitus; NYHA: New York Heart Association; LVEF: Left Ventricle Ejection Fraction; BB: Beta-Blockers; RAAS: Renin-Angiotensin-Aldosterone System; TTR: Transthyretin; MACE: Major Adverse Cardiovascular Events; Afib: Atrial Fibrillation; VT: Ventricular Tachycardia; CV: Cardiovascular; HFF: Heart Failure Hospitalization.

### 2.4 Quality and GRADE Assessments

Studies were evaluated using the Risk of Bias in Nonrandomized Studies of Interventions (ROBINS-I) tool, which evaluates studies in seven different domains categorizing them as “low risk”, “moderate risk”, “serious risk” and “critical risk” of bias.^12^ Additionally, we assessed publication bias for the primary outcome through funnel plot analysis. We evaluated the certainty of existing evidence by implementing the GRADE (Grading of Recommendations, Assessment, Development, and Evaluations) approach for the primary outcome.^13^ The certainty of the evidence was initially appraised as ‘high’, and was downgraded to ‘moderate’, ‘low’, or ‘very low’ by assessing studies based on five domains: study limitations, indirectness, imprecision, inconsistency, and publication bias.

### 2.5 Statistical Analysis

Risk ratios (RR) with 95% confidence intervals (CI) were calculated to evaluate treatment effects for binary outcomes, with statistical significance set at a p-value < 0.05. A Mantel-Haenszel (MH) test with a random-effects model was employed based on the level of heterogeneity for each outcome, and results were visually represented using a forest plot. Study heterogeneity was assessed through the Cochran Q test and the I² statistic. We followed the Cochrane pre-defined thresholds for heterogeneity interpretation: 0–40% indicating minimal heterogeneity, 30–60% moderate, 50–90% substantial, and 75–100% considerable heterogeneity. Additionally, to assess the reliability and robustness of the findings as well as explore possible causes of heterogeneity, we performed a sensitivity analysis using the leave-one-out strategy. RStudio version 4.2.2 (R Foundation for Statistical Computing) was used for all statistical analyses.

To better assess potential type I and type II errors, we performed a trial sequential analysis (TSA) for all outcomes. We used a random-effects model with 95% CI, an information axis with sample size, type one error with 5% two-sided boundary, and power of 80%. The adjustment of the thresholds for the Z score was based on the O’Brien–Fleming alpha spending function. TSA was performed using the TSA program version 0.9.5.10 beta (Copenhagen Trial Unit, Centre for Clinical Intervention Research, Rigshospitalet, Copenhagen, Denmark).^14^

### 2.6 Data Sharing Statement

No additional data are available.

## 3. Results

### 3.1 Study selection and characteristics

As shown in Figure 1, our initial search identified 188 records. After removing 84 duplicates and excluding irrelevant studies, 16 articles were fully evaluated for eligibility criteria. Of these, six were excluded due to the absence of a control group, four were eliminated because of overlapping populations, and two were omitted for not reporting the outcome of interest. Ultimately, four observational studies encompassing a total of 5039 patients were included in this systematic review and meta-analysis. Among these patients, 2489 (49.39%) received SGLT2 inhibitors, while 2550 (50.61%) were SGLT2 inhibitors-naïve. The patient cohort was predominantly male (3419, 67.85%), with a mean age of 78.25 years and an average LVEF of 50%. Additionally, 2573 (51.06%) patients had diabetes mellitus, and the mean follow-up duration ranged from 26 to 36 months. Study characteristics are detailed in Table 1.

**Figure 1.**
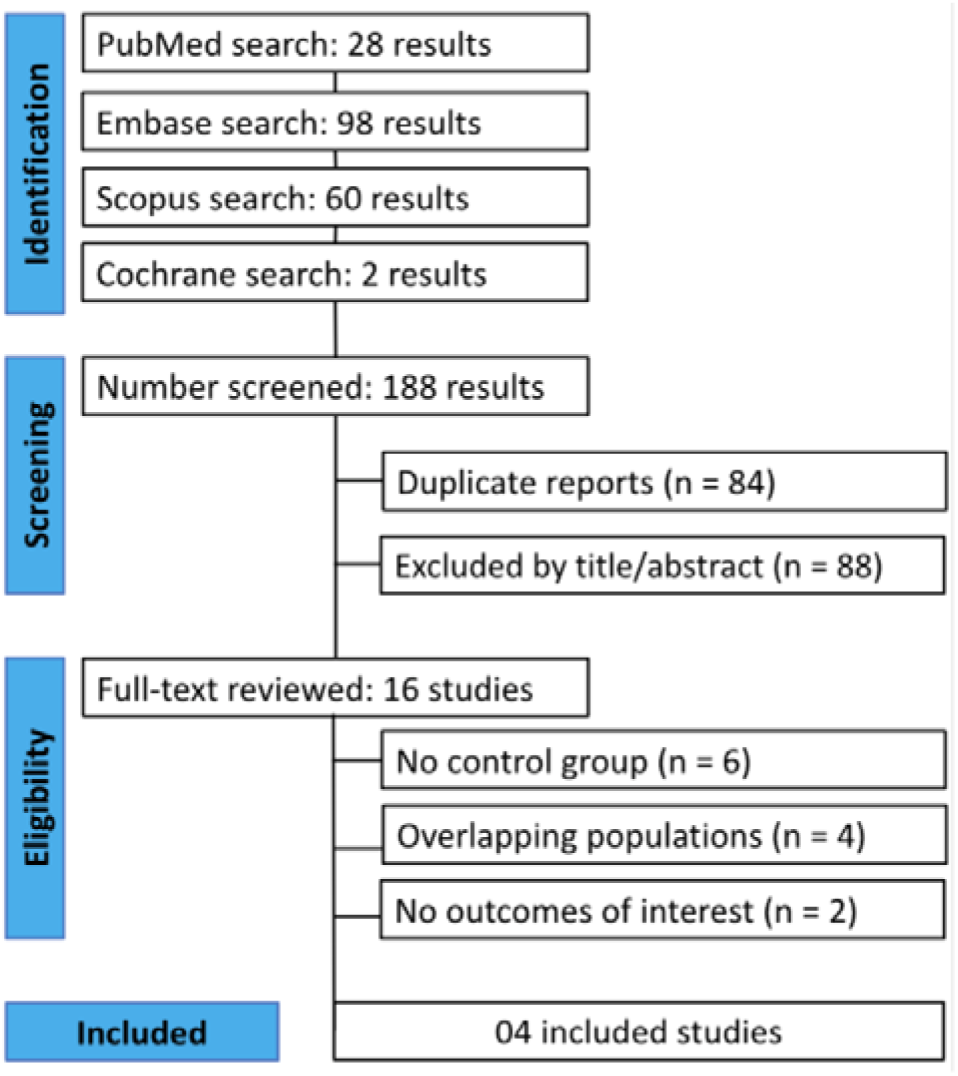
PRISMA flow diagram of study screening and selection.

### 3.2 Qualitative synthesis

All four included studies were observational in nature, three, Gonzalez-Montes et al., Schwegel et al., and Porcari et al., were prospective cohort studies, and one, Jaiswal et al., was retrospective. In terms of SGLT2 inhibitors evaluated, all studies included dapagliflozin and empagliflozin. Porcari et al. also examined canagliflozin, and Jaiswal et al. treated SGLT2i use as a class effect, without differentiating between specific agents. Notably, none of the studies applied exclusion criteria based on the type of SGLT2i prescribed. Consequently, the predominance of one agent over another likely reflects real-world prescribing patterns rather than methodological bias, which supports interpreting the results from a class-effect perspective rather than as head-to-head comparisons. All-cause mortality was consistently reported across studies, providing a common primary endpoint. However, secondary outcomes varied. Jaiswal et al. evaluated a broad range of CV outcomes, including major adverse cardiovascular events (MACE), ischemic stroke, heart failure, atrial fibrillation, and ventricular tachycardia, while Porcari et al. and Schwegel et al. focused more specifically on CV death and hospitalizations due to worsening heart failure (WHF). Gonzalez-Montes et al. uniquely reported adverse events alongside mortality. Despite differences in outcome scope and design, all studies assessed the long-term impact of SGLT2 inhibitors in routine clinical practice, with follow-up durations ranging from just over two years to three years. These shared features support the generalizability of findings, while variations in methodology, outcome definitions, and drug exposure contribute to the clinical and methodological heterogeneity that must be carefully considered when interpreting pooled results.

### 3.3 Pooled analysis of all studies

In the SGLT2 inhibitors group (n=2489), 335 all-cause deaths were recorded compared to 688 in the control group (n=2550). Analysis revealed that all-cause mortality (RR 0.44; 95% CI 0.33-0.59; p<0.00001; I²=54%; Figure 2) was significantly lower in patients treated with SGLT2 inhibitors, corresponding to a 56% reduction in risk relative to controls. Similarly, 21 CV deaths occurred in the SGLT2 inhibitors group (271), contrasting with the 78 deaths reported in the control group (285); meaning CV mortality (RR 0.30; 95% CI 0.16-0.55; p=0.0001; I²=25%; Figure 3) was significantly lower among patients receiving SGLT2 inhibitors. In contrast, although there was a downward trend in heart failure hospitalizations (RR 0.68; 95% CI 0.33-1.41; p=0.30; I²=89%; Figure 4), this result did not reach statistical significance.

**Figure 2.**
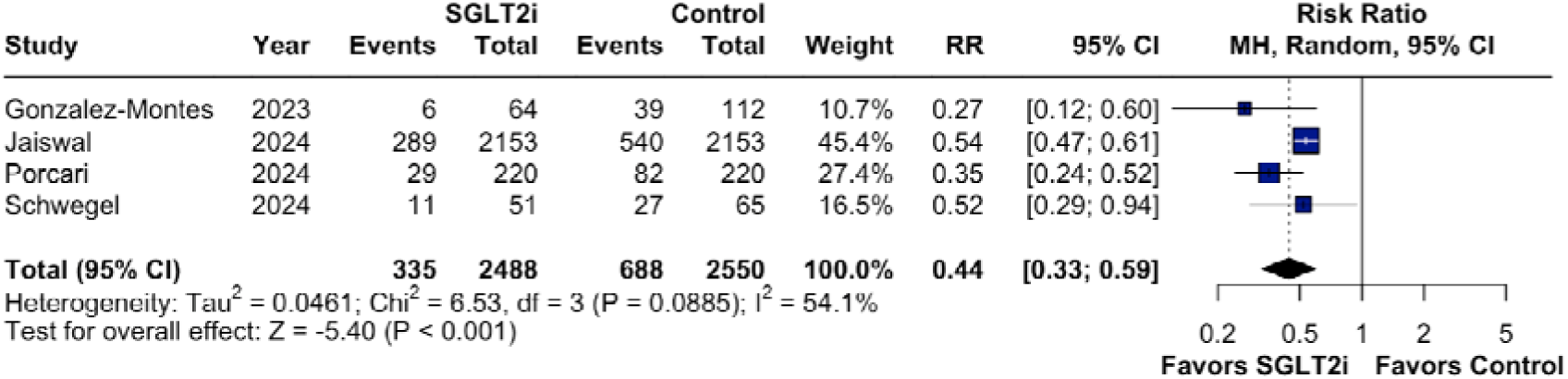
Significantly lower all-cause mortality in patients treated with SGLT2 inhibitors.

**Figure 3.**
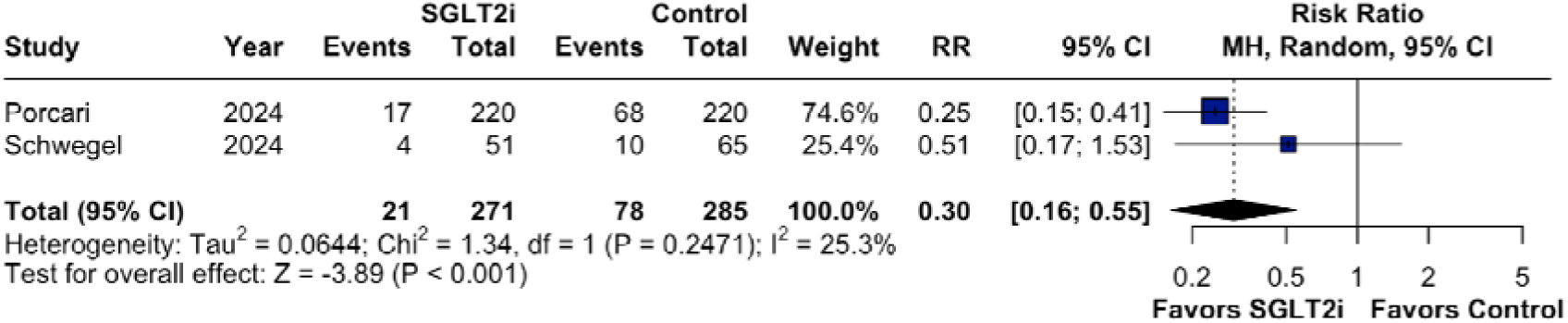
Statistically significant lower cardiovascular mortality among patients receiving SGLT2 inhibitors compared to the control group.

**Figure 4.**
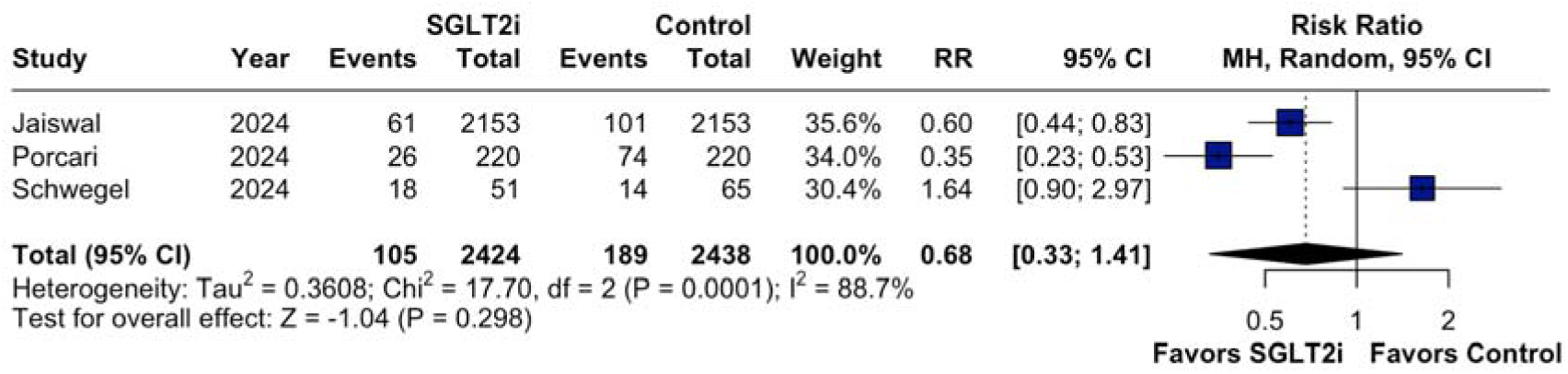
No statistically significant difference in heart failure hospitalizations between the analyzed groups.

### 3.4 Risk of Bias in Included Studies and GRADE Assessment

Quality assessment of the included studies was performed independently by M.C. and V.L. Discrepancies were resolved through discussion and re-evaluation. One study was categorized as having a serious risk of bias due to the authors not using an analysis method that controlled for important confounders. The remaining studies were deemed to have a moderate risk of bias, as the authors employed propensity score matching or other analytical methods to account for potential confounders (Supplemental Table 1). Additionally, there was no indication of publication bias, as the funnel plot analysis revealed a symmetrical distribution of studies with comparable weights around the meta-analysis point estimate (Supplemental Figure 1). Furthermore, we evaluated the certainty of existing evidence by implementing the GRADE approach for all outcomes. (Supplemental Table 2).

### 3.5 Sensitivity Analysis

A leave-one-out sensitivity analysis was performed to evaluate the robustness of the findings for the all-cause mortality outcome. The RR remained consistent across all iterations, ranging from 0.37 to 0.49, with all estimates retaining statistical significance. The overall pooled RR was 0.44 [95% CI: 0.33 to 0.59], with moderate heterogeneity (I² = 54.1%) (Supplementary Figure 2). These results indicate that no single study disproportionately influenced the overall effect size, underscoring the stability and reliability of the observed mortality benefit associated with SGLT2 inhibitor use in patients with ATTR-CM.

For cardiovascular mortality, the leave-one-out analysis revealed some sensitivity to the inclusion of specific studies. While the overall pooled estimate remained statistically significant (RR: 0.30 [95% CI: 0.16 to 0.55], I² = 25.3%) (Supplementary Figure 3), omission of the Schwegel et al. study further strengthened the effect (RR: 0.25 [95% CI: 0.15 to 0.41]). In contrast, excluding the Porcari et al. study resulted in a non-significant estimate (RR: 0.51 [95% CI: 0.17 to 1.53]), suggesting that the overall finding may be influenced by its inclusion.

In the analysis of HF hospitalizations, the pooled estimate was not statistically significant (RR: 0.68 [95% CI: 0.33 to 1.41]), and heterogeneity was substantial (I² = 88.7%) (Supplementary Figure 4). Exclusion of the Schwegel study yielded a significant effect (RR: 0.47 [95% CI: 0.28 to 0.80]), whereas omitting either Jaiswal or Porcari produced attenuated, non-significant estimates (RR: 0.75 [0.16 to 3.40] and RR: 0.96 [0.36 to 2.57], respectively). These findings suggest that the observed effect on HF hospitalizations lacks robustness and is particularly sensitive to the inclusion of individual studies.

### 3.6 Trial Sequential Analysis

The TSA for all-cause mortality showed that the cumulative Z-score crossed the conventional boundary of two, indicating statistically significant evidence favoring the use of SGLT2 inhibitors in reducing this outcome among patients with ATTR-CM. Moreover, the required information size of 1,720 was met, confirming the robustness of the finding. This suggests the presence of a true effect and implies that further trials may not be necessary to confirm these results (Supplemental Figure 5).

The TSA for CV-mortality indicated that the first information fraction exceeded 100% of the required information size, preventing the rendering of formal sequential monitoring boundaries. Nonetheless, the cumulative Z-score crossed the conventional significance threshold (Z > 2), suggesting statistically significant evidence of benefit. Given that the required information size was surpassed, these findings support the presence of a true effect; however, due to the inability to construct trial sequential monitoring boundaries, caution is advised in interpreting the conclusiveness of these results. (Supplemental Figure 6).

The TSA for HF-hospitalizations demonstrated that neither the required information size nor the conventional significance threshold (Z = 2) was reached. This suggests that the available data are underpowered and inconclusive, indicating a need for further high-quality randomized controlled trials to establish the effect of SGLT2 inhibitors on this outcome in patients with ATTR-CM. (Supplemental Figure 7).

## Discussion

In this systematic review and meta-analysis of four observational studies (2 with propensity score-matched (PSM) cohorts) and 5039 patients, we compared therapy with SGLT2 inhibitors to SGLT2 inhibitors-naïve patients with ATTR-CM. The key findings of our study are as follows: (1) Patients treated with SGLT2 inhibitors experienced a 56% reduction in all-cause mortality compared to those who were SGLT2 inhibitor-naïve. (2) SGLT2 inhibitor therapy was associated with a 70% decrease in CV mortality. (3) While there was a trend toward fewer HF hospitalizations in the SGLT2 inhibitor group, this reduction was not statistically significant.

A recent study by Ioannou et. al., reported no relevant effect of treatment with angiotensin inhibitors (ACEis/ARBs) and beta-blockers in a cohort of 2371 patients with ATTR-CM.^15^ Additionally, several authors have reported the poor tolerability of standard HF medications in this specific population. The infiltrative nature of ATTR-CM leads to myocardial stiffening and diastolic dysfunction, limiting cardiac reserve and making patients more susceptible to hemodynamic disturbances.^6^ As a result, such agents which may precipitate significant hypotension are frequently challenging to use in this population—and their effects are uncertain.^15^ Consequently, clinicians often face difficulties in initiating and titrating conventional HF therapies in ATTR-CM patients, sometimes resulting in suboptimal dosing or complete avoidance of these medications; and therefore relying more on supportive care such as cautious diuretic use.^6, 8^ These factors, combined with the limited use of the only two FDA-approved disease-modifying therapies, create a scenario in which therapeutic options for most ATTR-CM patients remain scarce.

ATTR-CM is increasingly recognized as a frequent cause of HFpEF, as evidenced by studies from Gonzales-Lopez et al. and Mohammed et al., who detected ATTR amyloid in 13–19% of patients with HFpEF.^16–17^ Because HF trials did not implement rigorous screening to exclude cardiac ATTR amyloidosis, it is possible that some patients with undiagnosed ATTR amyloidosis were included. This may partly explain the negative outcomes observed in previous studies on HFpEF treatment.^6^ In recent years, SGLT2 inhibitors have transformed HF treatment, with guidelines now endorsing their use across all types and stages of HF.^6–7^ Specifically, the European Society of Cardiology (ESC) has given a Class IA recommendation for SGLT2 inhibitors in patients with HFpEF, aiming to reduce the risk of HF hospitalization and CV death.^18^ Moreover, preliminary observational studies have demonstrated that SGLT2 inhibitors are safe and well tolerated in patients with ATTR-CM, suggesting their potential as a novel therapeutic option for this patient population; primarily due to their capacity to improve fluid balance without compromising the systemic blood pressure.^19–21^ Studies by Lang et al. and Dobner et al., demonstrated that SGLT2 inhibitors are generally well tolerated, without an increase in typical adverse events despite a theoretical increased risk (driven by advanced age, autonomic neuropathy, prostatic hypertrophy and often poor nutritional status), and a favorable clinical and cardiac-metabolic profile similar to the one observed in other non-ATTR clinical studies such as EMPEROR-Preserved.^21–22^

Our findings are consistent with those reported in most individual studies, reinforcing the potential role of SGLT2 inhibitors in the treatment strategy for ATTR-CM. For example, Porcari et al. and Schwegel et al. independently found that treatment with SGLT2 inhibitors was significantly associated with lower all-cause mortality, CV mortality, and reduced rates of HF hospitalization-observations that held true regardless of variations in renal function, NT-proBNP levels, systolic function, or the use of concomitant tafamidis.^2, 23^ Similarly, Jaiswal et al. confirmed these outcomes in a larger and more diverse cohort during both short- and long-term follow-up, also reporting improvements in other endpoints such as ischemic stroke and atrial fibrillation in patients treated with SGLT2 inhibitors.^24^

Our study has several limitations that must be acknowledged. First, all included studies were observational, and none were randomized controlled trials (RCTs), which inherently increases the risk of bias. Although two of the four studies employed propensity score matching to mitigate confounding, this approach does not fully replicate the balance achieved through randomization, leaving residual confounding as a concern. Additionally, indication bias may be present-clinicians might have preferentially prescribed SGLT2 inhibitors to patients with either more advanced disease or, conversely, to those perceived as lower risk due to the lack of robust data in this population. Heterogeneity in study design, patient characteristics, and outcome definitions-particularly in the case of heart failure hospitalizations-may have also influenced the results. Moreover, our study was underpowered to conduct meaningful subgroup analyses, limiting the ability to explore differential treatment effects across specific patient populations. Taken together, while our findings are encouraging, they highlight the urgent need for large-scale, prospective RCTs to definitively establish the role of SGLT2 inhibitors in the management of ATTR-CM.

## Conclusions

This meta-analysis, incorporating data from 5,039 patients with ATTR-CM, provides evidence supporting the potential benefit of SGLT2 inhibitors in this population. Treatment with SGLT2 inhibitors was associated with a statistically significant reduction in both all-cause and CV mortality. While a favorable trend toward reduced HF hospitalizations was observed, this finding did not reach statistical significance and demonstrated substantial heterogeneity.

Leave-one-out sensitivity analyses confirmed the robustness of the all-cause mortality benefit, with consistent and statistically significant results across all iterations. In contrast, the effects on CV mortality and HF hospitalizations were more sensitive to the inclusion or exclusion of individual studies, suggesting these findings may be less stable. Furthermore, trial sequential analysis indicated that the available evidence for non-mortality outcomes remains insufficient to draw definitive conclusions. In contrast, the strength of the all-cause mortality finding-both in sensitivity and trial sequential analysis-supports its reliability and underscores its potential clinical relevance.

Although these findings align with emerging data from other populations, the observational design and methodological limitations of the included studies such as potential residual confounding and selection bias highlight the urgent need for well-designed RCTs to validate the therapeutic role of SGLT2 inhibitors in patients with ATTR-CM.

## Data Availability

All data referred to in this manuscript are available within the published articles included in the systematic review and meta-analysis. Extracted datasets and analysis code used for pooling results are available from the corresponding author upon reasonable request.

## Abbreviations

ATTR-CM: Transthyretin amyloid cardiomyopathy
CI: Confidence interval
CV: Cardiovascular
HF: Heart failure
HFpEF: Heart failure with preserved ejection fraction
RCT: Randomized controlled trial
ROBINS-I: Risk of Bias in Nonrandomized Studies of Interventions
RR: Risk ratio
RRR: Relative risk reduction
SGLT2: Sodium-glucose cotransporter 2

---

**Supplemental Figure 1.**
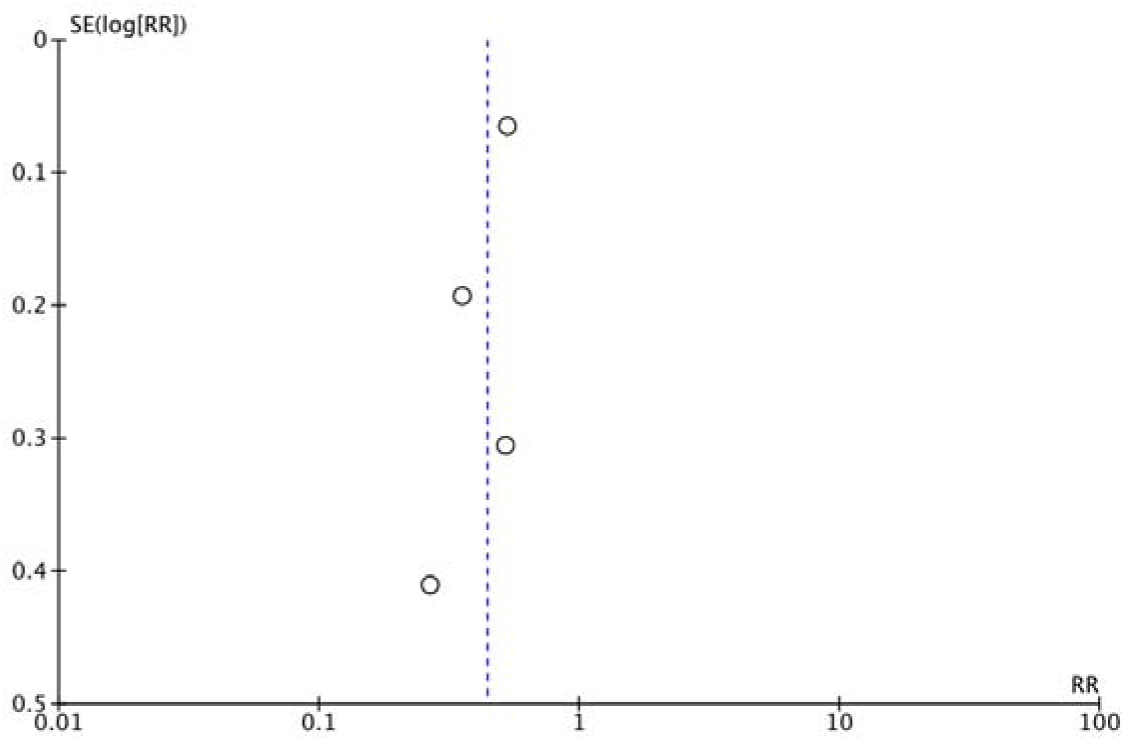
Funnel-plot analysis for all-cause mortality.

**Supplemental Figure 2.**
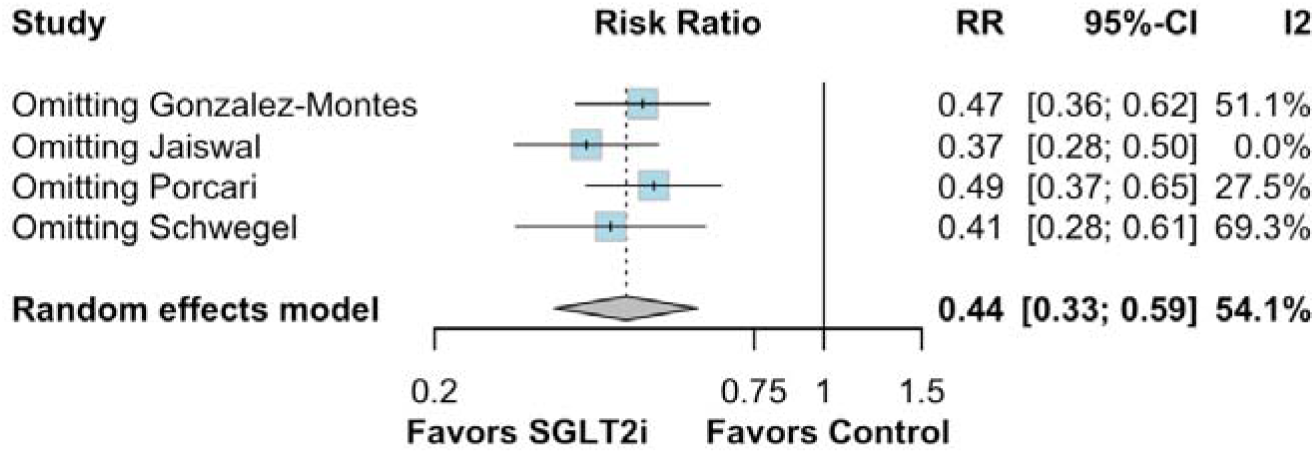
Leave-one-out sensitivity analysis for all-cause mortality.

**Supplemental Figure 3.**
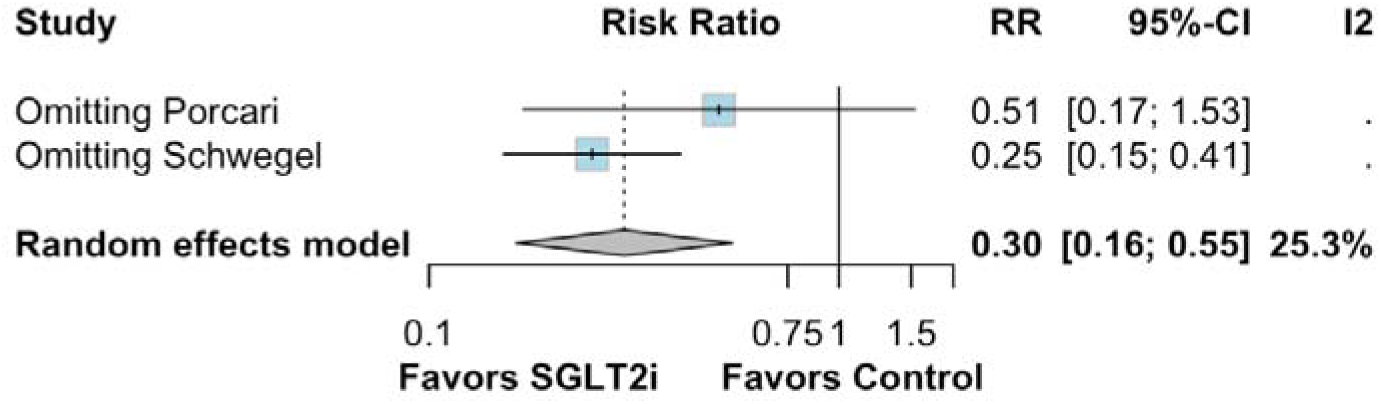
Leave-one-out sensitivity analysis for cardiovascular mortality.

**Supplemental Figure 4.**
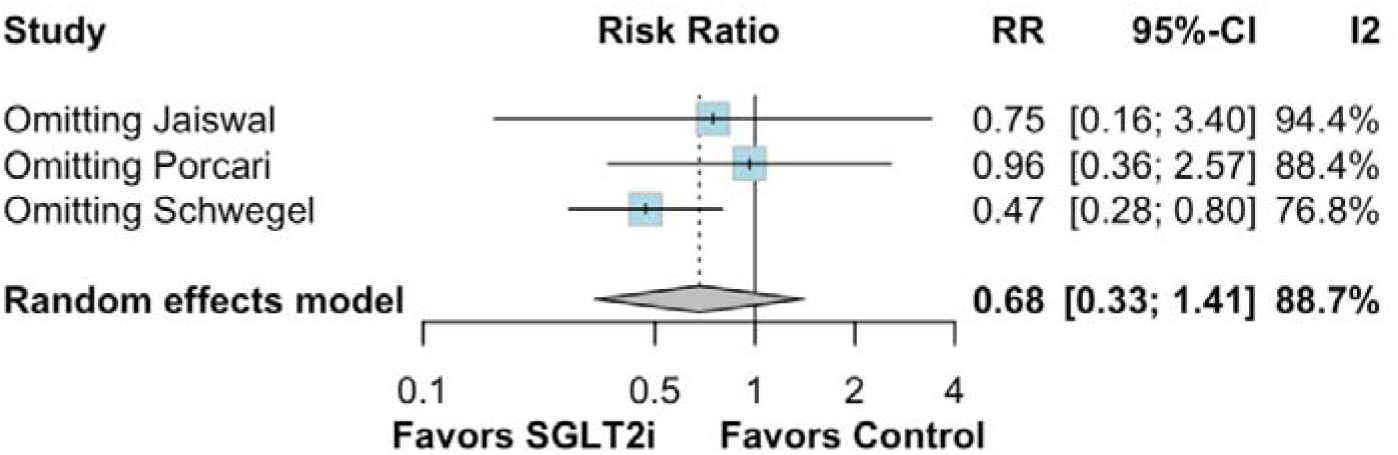
Leave-one-out sensitivity analysis for heart failure hospitalizations.

**Supplemental Figure 5.**
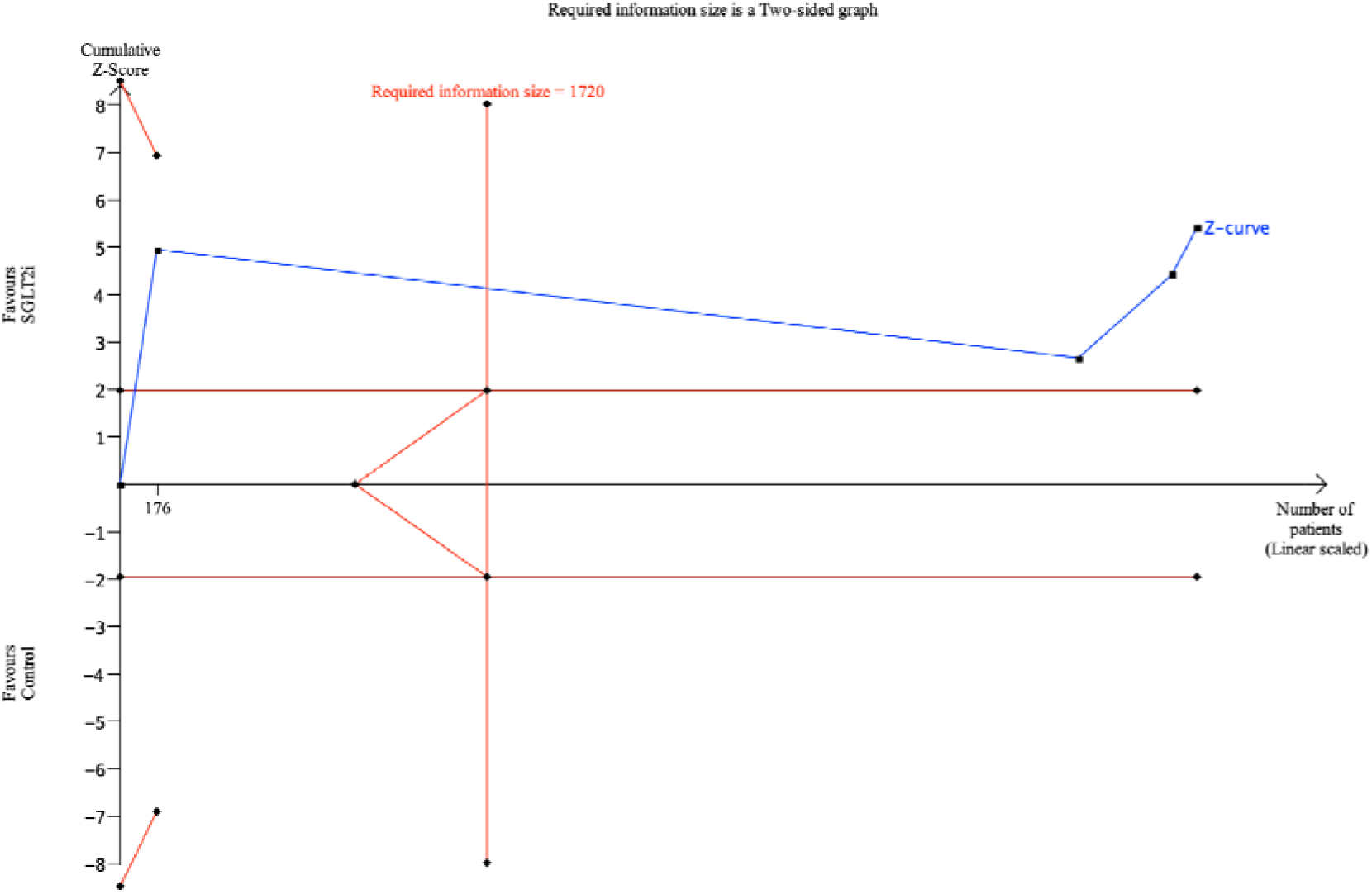
Trial sequential analysis for all-cause mortality with required information size achieved.

**Supplemental Figure 6.**
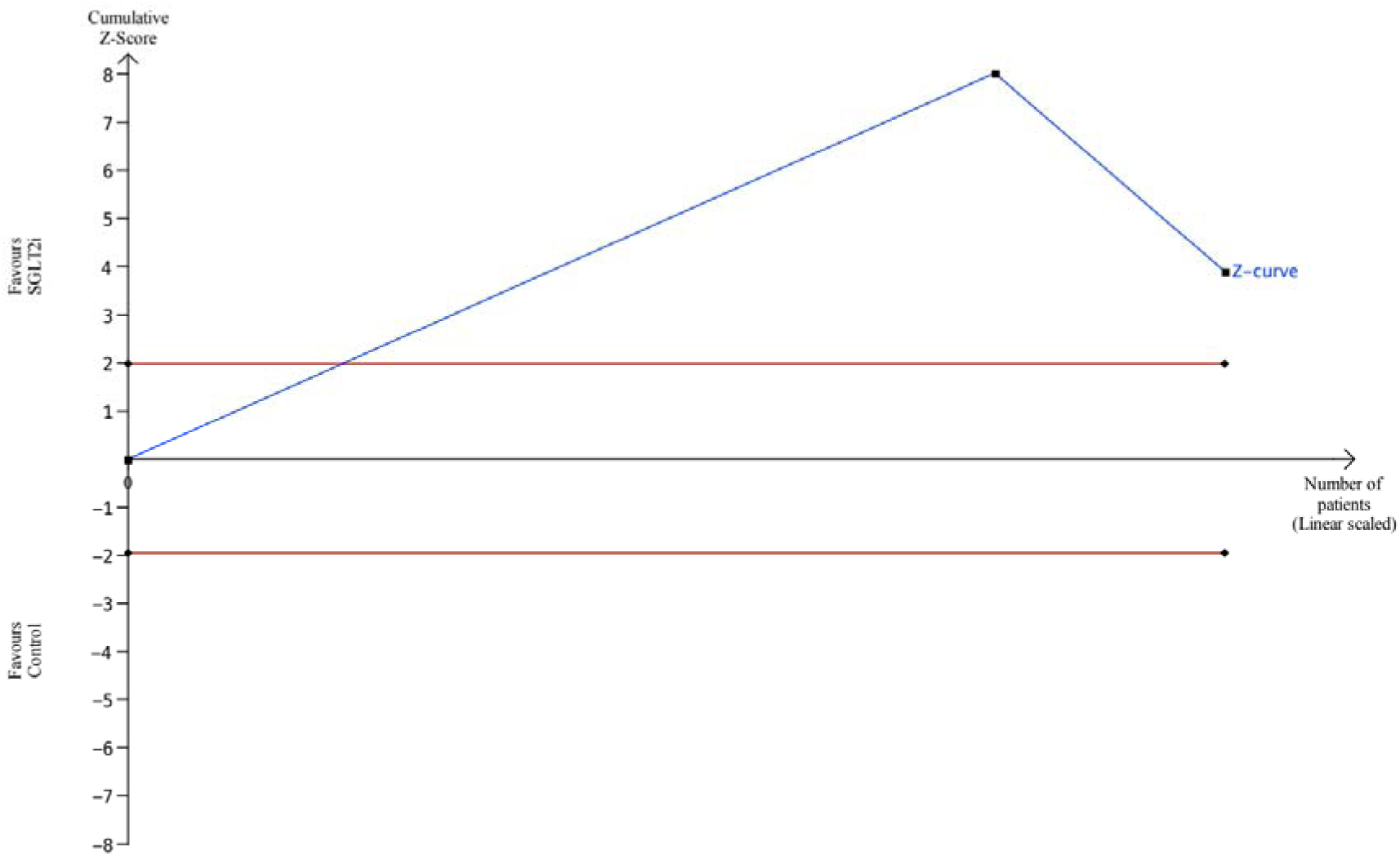
Trial sequential analysis for cardiovascular mortality with the information fraction exceeding 100% of the required information size.

**Supplemental Figure 7.**
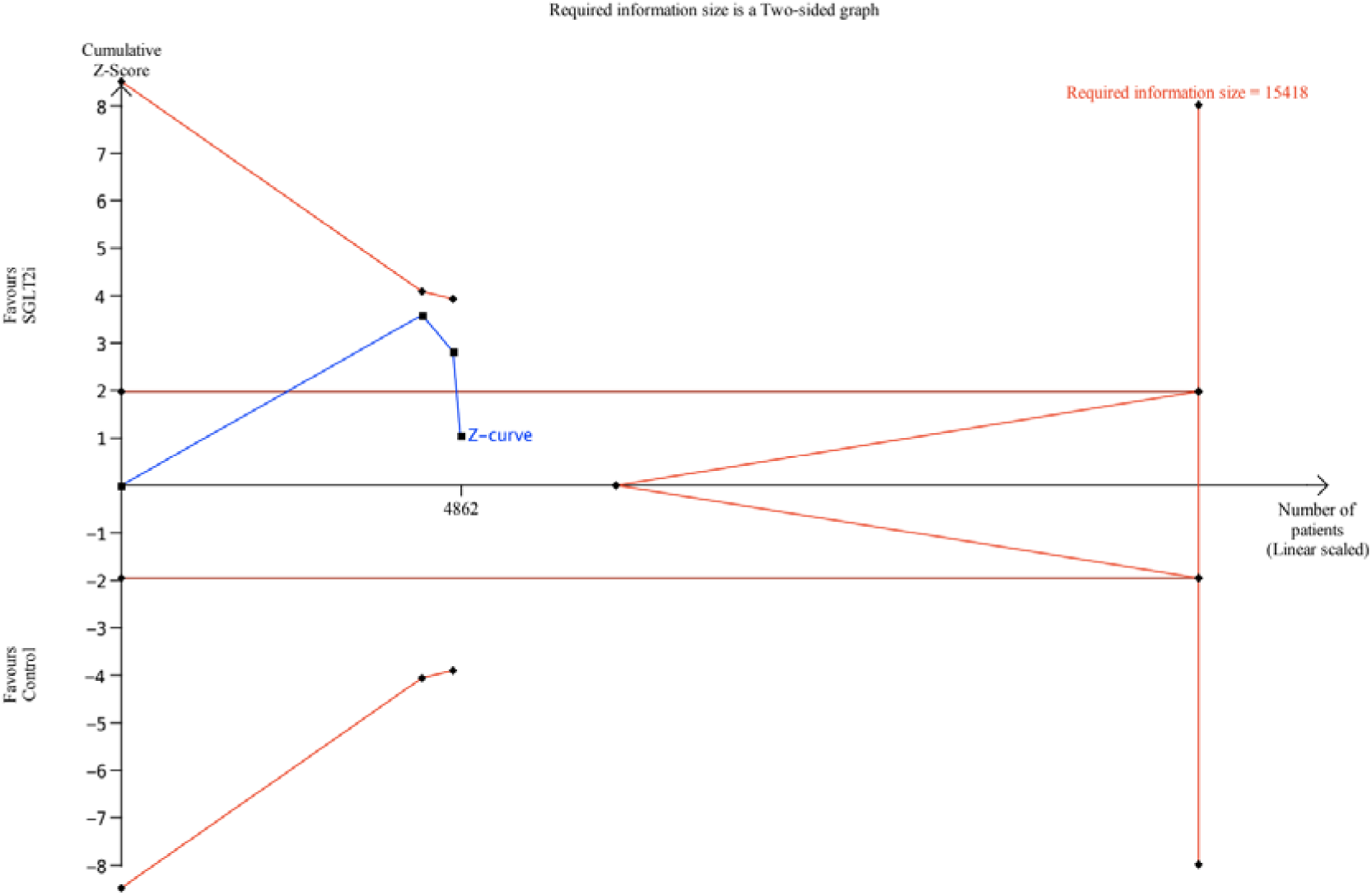
Trial sequential analysis for heart failure mortality with failure to achieve required information size.

**Supplemental Table 1.** ROBINS-I tool for risk of bias assessment.

| Study | Bias due to confounding | Bias in selection of participants into the study | Bias in classification of interventions | Bias due to deviations from intended interventions | Bias due to missing data | Bias in measurement of outcomes | Bias in selection of the reported result | Overall Bias |
| --- | --- | --- | --- | --- | --- | --- | --- | --- |
| Gonzalez-Montes 2023 | Serious | Low | Low | Low | Low | Low | Moderate | Serious |
| Jaiswal 2024 | Moderate | Low | Low | Low | Low | Low | Low | Moderate |
| Porcari 2024 | Moderate | Low | Low | Low | Low | Low | Low | Moderate |
| Schwege I 2024 | Moderate | Low | Low | Low | Low | Low | Low | Moderate |

**Supplemental Table 2.**
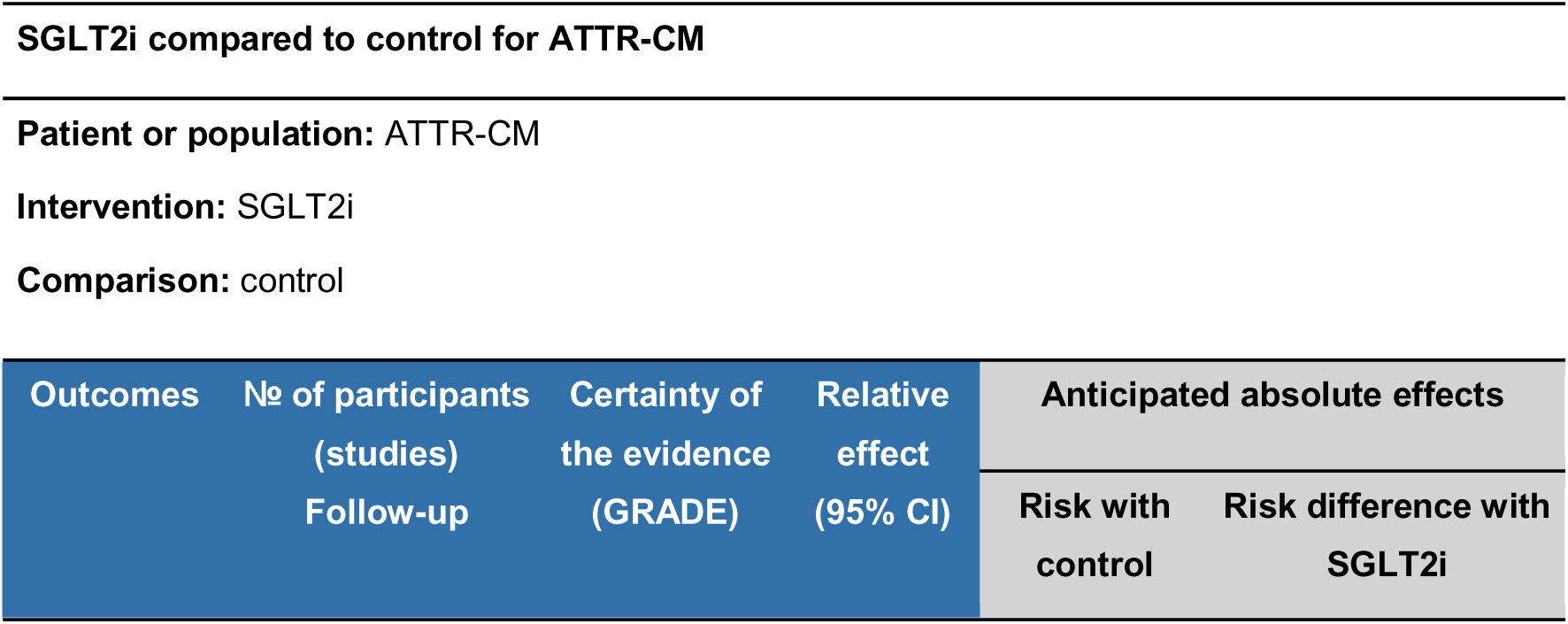
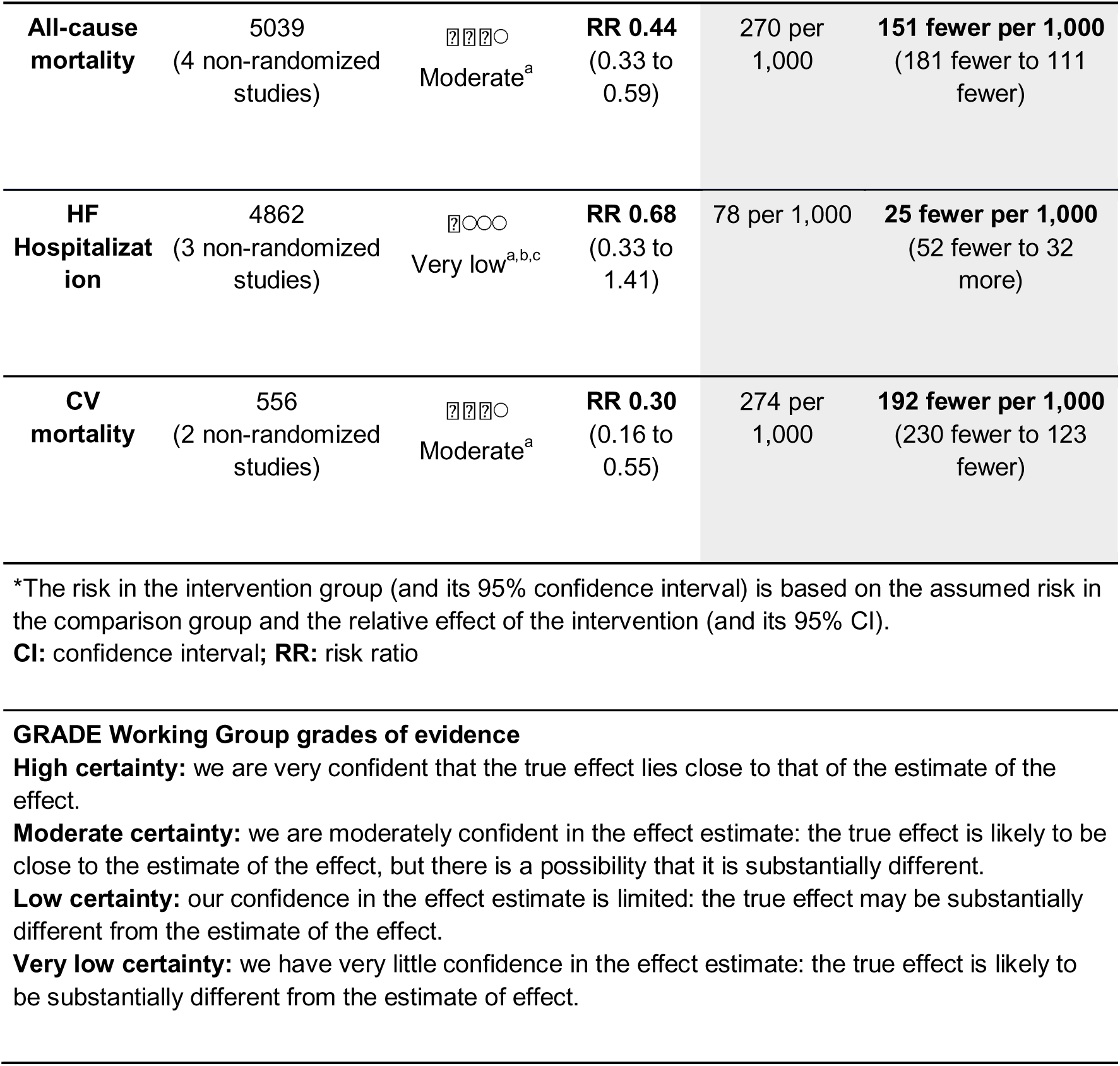
GRADE assessment results.

| SGLT2i compared to control for ATTR-CM |  |  |  |  |  |
| --- | --- | --- | --- | --- | --- |
| Patient or population: ATTR-CM |  |  |  |  |  |
| Intervention: SGLT2i |  |  |  |  |  |
| Comparison: control |  |  |  |  |  |
| Outcomes | No of participants (studies)<br>Follow-up | Certainty of the evidence (GRADE) | Relative effect (95% CI) | Anticipated absolute effects |  |
|  |  |  |  | Risk with control | Risk difference with SGLT2i |
| **All-cause mortality** | 5039<br>(4 non-randomized studies) | Moderate<sup>a</sup> | **RR 0.44**<br>(0.33 to 0.59) | 270 per 1,000 | **151 fewer per 1,000**<br>(181 fewer to 111 fewer) |
| **HF Hospitalization** | 4862<br>(3 non-randomized studies) | Very low<sup>a,b,c</sup> | **RR 0.68**<br>(0.33 to 1.41) | 78 per 1,000 | **25 fewer per 1,000**<br>(52 fewer to 32 more) |
| **CV mortality** | 556<br>(2 non-randomized studies) | Moderate<sup>a</sup> | **RR 0.30**<br>(0.16 to 0.55) | 274 per 1,000 | **192 fewer per 1,000**<br>(230 fewer to 123 fewer) |

\*The risk in the intervention group (and its 95% confidence interval) is based on the assumed risk in the comparison group and the relative effect of the intervention (and its 95% CI).
**CI:** confidence interval; **RR:** risk ratio

## Explanations

a. Outcome carried out with studies with moderate-serious risk of bias. b. High heterogeneity. c. Wide confidence interval (RR 0.68; 95% CI 0.33-1.41)

